# Factors Associated with Drug Interactions in Medical Prescriptions Received in Community Pharmacies in Yaoundé

**DOI:** 10.1101/2025.06.25.25330261

**Authors:** Donald Embogo, C Nnanga, AP Minyem Ngombi, Sandra Elono B., Eric Nseme

## Abstract

**Introduction:** Prescriptions involving multiple drugs issued by healthcare professionals are frequently at the origin of drug–drug interactions. These interactions represent a major cause of poor adherence, therapeutic failure, or potentially serious adverse events that may compromise the patient’s vital prognosis. In Cameroon, drug interactions have been studied on several occasions in hospital settings but only rarely in community pharmacies. This motivated our study, which aimed to analyse the risk factors associated with the potential occurrence of drug interactions in medical prescriptions received in pharmacies within the Efoulan Health District in Yaoundé.

**Methodology:** A descriptive cross-sectional study with prospective data collection was conducted over a 7-month period, from November 2022 to May 2023, in all pharmacies within the Efoulan Health District. All legible and compliant prescriptions containing at least two medications were included. Data were collected using a pre-designed and pre-tested data collection form. Drug interactions were identified using the THERIAQUE® drug database. Data analysis was performed using IBM-SPSS® Version 23.0.

**Results:** The study included 385 prescriptions, with a frequency of drug interactions in medical prescriptions of **17.5%**. The prescriptions received were mainly issued by general practitioners (**43.9%**). The median number of drugs prescribed per prescription was **2 [2–3]**, with extremes ranging from 2 to 6. Most prescriptions contained two medications (**42.1%**). Prescriptions with two or three drugs, as well as those with more than three drugs, were predominantly written by general practitioners, accounting for **24.9%** and **19%**, respectively. However, cardiologists were twice as likely as other prescribers to prescribe more than three drugs. It was statistically demonstrated that the greater the number of prescribed medications, the higher the likelihood of drug interactions. Neurologists and cardiologists were, respectively, **7 times** and **6 times** more likely to issue prescriptions involving drug–drug interactions than other prescribers.

**Conclusion:** Drug interactions represent a moderate proportion of medical prescriptions received in community pharmacies in the Efoulan Health District of Yaoundé. They are statistically associated with the number of medications prescribed and with prescriptions initiated by certain specialists, notably neurologists and cardiologists.

## INTRODUCTION

A medical prescription is a clinical act that involves recording a treatment on a document: the prescription form, which serves as the main tool of communication between the prescriber and the pharmacist [1]. From a regulatory standpoint, a medical prescription constitutes the culmination of a structured process and is considered a medico-legal act that engages both the civil and criminal liability of the healthcare professional who writes it and the pharmacist who dispenses it [2].

Prescribed medicines may interact with each other, with food, or with dietary supplements, giving rise to drug–drug interactions (DDIs). These are defined as pharmacological, pharmacokinetic or clinical responses resulting from a modification—either a reduction or an enhancement—of the effect of a drug due to the prior or concomitant administration of another substance [3]. DDIs are a major cause of poor adherence, therapeutic failure, or potentially serious adverse events that may compromise the patient’s prognosis, particularly in the context of polypharmacy [4]. It has been estimated that 42% of adverse drug events are preventable, most of which occur at the prescription stage, and a large proportion of these are due to drug interactions [5]. Such interactions may occur in both hospital settings and community pharmacies.

While several studies worldwide have examined drug interactions in hospitals, relatively few have addressed them in the context of community pharmacies. In Cameroon, a study conducted in 2021 at Jamot Hospital in Yaoundé found that nearly 80% of psychotropic drug prescriptions contained at least three drug interactions [6]. In an effort to shed light on this essential issue, we undertook this study to analyse the risk factors associated with the likelihood of drug interactions in medical prescriptions received in community pharmacies in the Efoulan Health District, Yaoundé.

## METHODOLOGY

A descriptive cross-sectional study with prospective data collection was conducted in all community pharmacies located within the Efoulan Health District of Yaoundé over a 7-month period, from 1st November 2022 to 25th May 2023. The study population consisted of all medical prescriptions received in pharmacies within the Efoulan Health District of Yaoundé.

Inclusion criteria comprised all prescriptions containing at least two medications, with identifiable patient and prescriber information, and for which signed informed consent was obtained from the patient. Prescriptions that were illegible were excluded.

Data were collected using an anonymous, pre-designed and pre-tested data collection form. Drug interactions listed on prescriptions were identified using the THERIAQUE® drug database, a comprehensive database of all medicines available in France, designed for use by healthcare professionals.

Data entry and analysis were carried out using SPSS software, version 23.0. Qualitative variables were presented as counts and frequencies, while quantitative variables were summarised using median and standard deviation. Associations between qualitative variables were assessed using the Chi-square test, with a significance threshold set at p ≤ 0.05.

## RESULTS

### Frequency of Drug Interactions

The frequency of drug–drug interactions was 17.5% of the prescriptions studied, corresponding to 67 cases.

### Professional Qualifications of Prescribers

With regard to prescriber qualifications, the majority were general practitioners (43.9%), followed by specialists such as gynaecologists (13.0%), cardiologists (7.0%), and ophthalmologists (7.0%), as shown in Table I below.

**Table I:** Professional qualifications of prescribers.

| Prescriber Qualification | Number (N=385) | Percentage (%) |
| --- | --- | --- |
| General Practitioner | 169 | 43.9 |
| Gynaecologist | 50 | 13.0 |
| Cardiologist | 27 | 7.0 |
| Ophthalmologist | 27 | 7.0 |
| Paediatrician | 21 | 5.5 |
| Nurse/Midwife | 19 | 4.9 |
| Surgeon | 12 | 3.1 |
| Dental Practitioner | 10 | 2.6 |
| ENT Specialist | 8 | 2.1 |
| Hepato-Gastroenterologist | 7 | 1.8 |
| Rheumatologist | 7 | 1.8 |
| Infectious Disease Specialist | 7 | 1.8 |
| Dermatologist | 6 | 1.6 |
| Neurologist | 5 | 1.3 |
| Others | 10 | 2.6 |

### Number of Drugs Prescribed

The median number of drugs prescribed per prescription was 3 [2–3], with a range from 2 to 6 drugs. Most prescriptions contained 2 drugs (42.1%), as illustrated in Figure 1 below.

**Figure 1:**
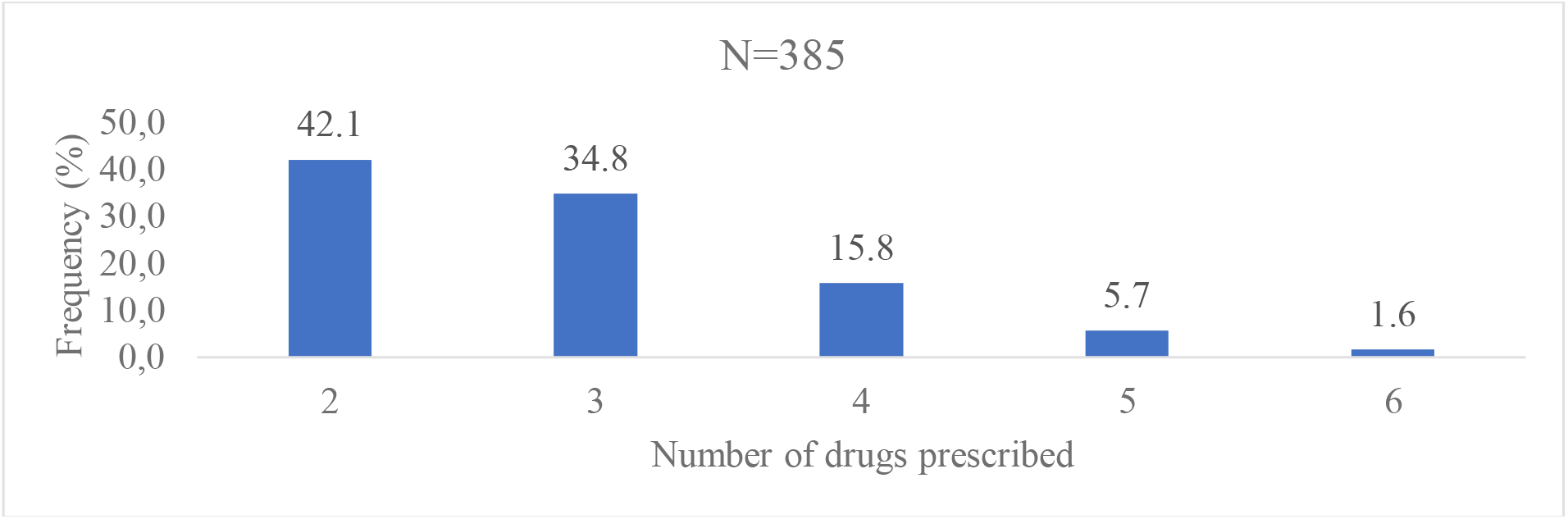
Distribution of prescriptions according to the number of drugs prescribed.

### Association Between Number of Drugs Prescribed and Prescriber Qualification

A significant association was found between the number of drugs prescribed and the qualification of the prescriber. In particular, cardiologists were twice as likely to prescribe three or more medications (OR: 2.70 [1.07–6.86]) as shown in Table II.

**Table II:**
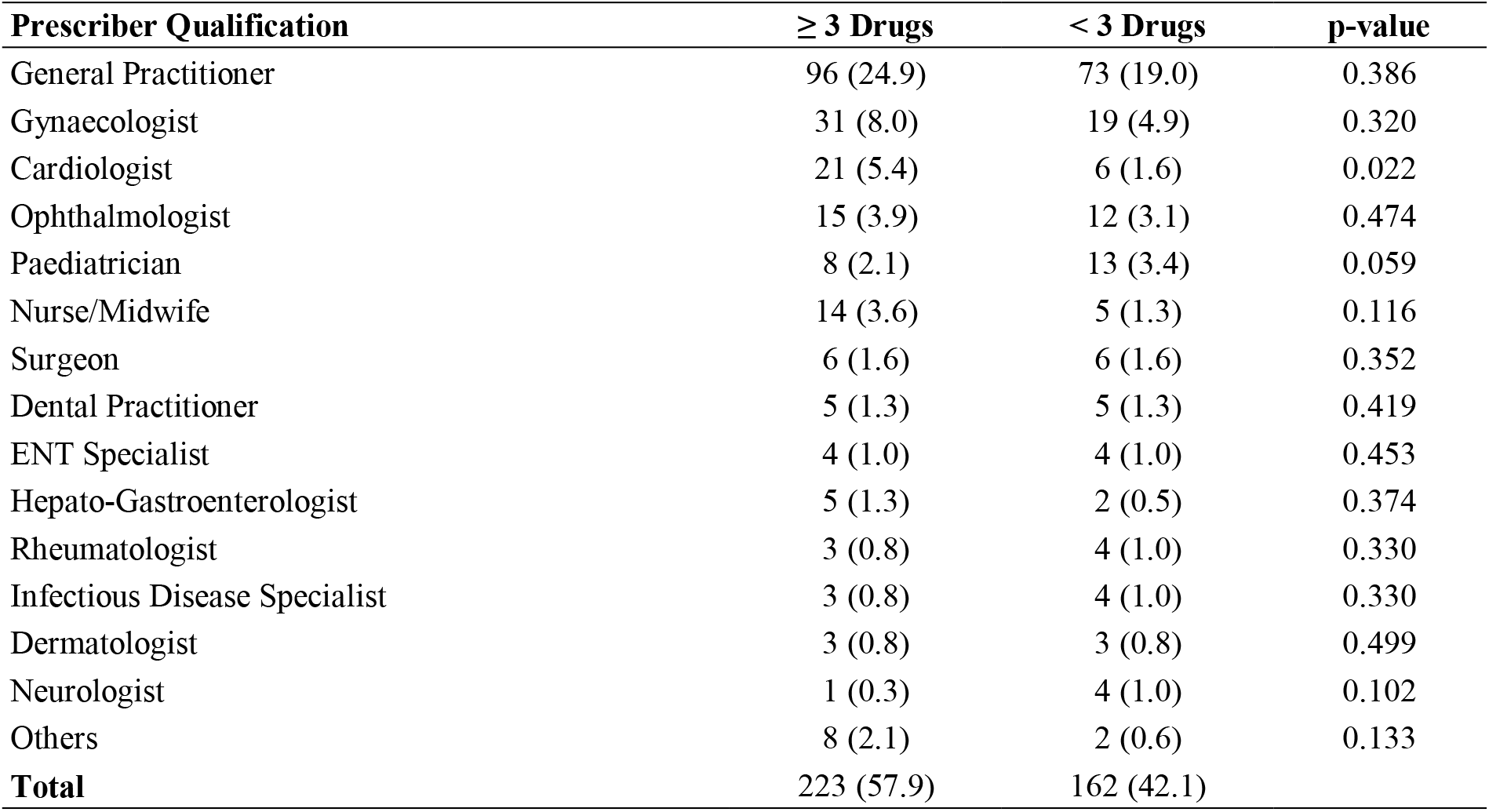
Association between number of drugs prescribed and prescriber qualification.

### Association Between Drug Interactions and Number of Drugs Prescribed

Table III demonstrates a statistically significant trend: the higher the number of drugs prescribed, the greater the likelihood of drug–drug interactions (p < 0.001).

**Table III:** Association between drug interactions and number of drugs prescribed.

| Number of Drugs Prescribed | With Interaction | Without Interaction | p-value |
| --- | --- | --- | --- |
| 2 | 10 (2.6) | 152 (39.5) |  |
| 3 | 23 (6.0) | 111 (28.8) |  |
| 4 | 15 (3.9) | 46 (11.9) |  |
| 5 | 13 (3.4) | 9 (2.3) |  |
| 6 | 6 (1.6) | 0 (0.0) | **<0.001** |
| **Total** | 67 (17.5) | 318 (82.5) |  |

**Table IV:**
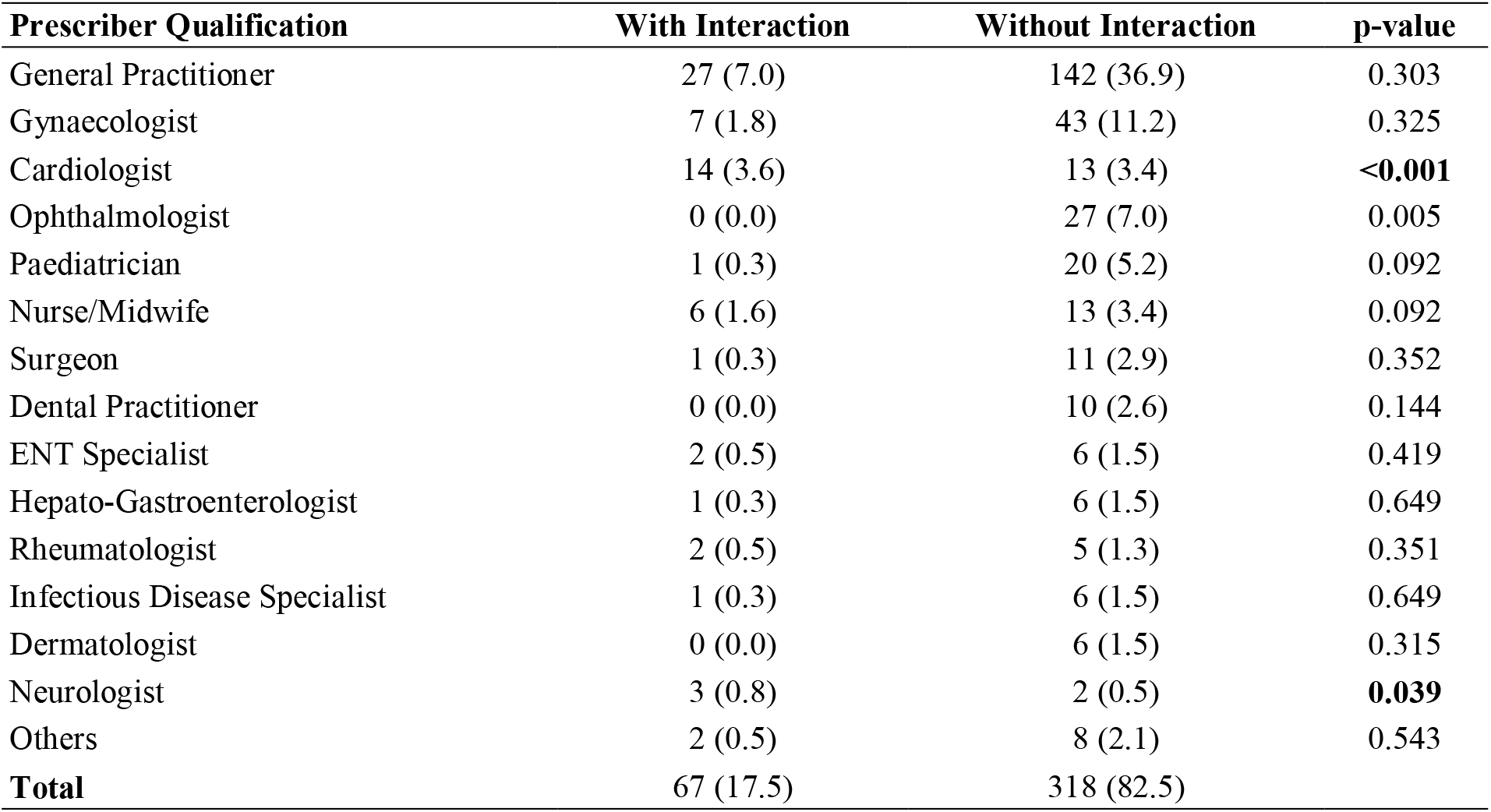
Association between prescriber qualification and occurrence of drug–drug interactions.

### Association Between Prescriber Qualification and Drug Interactions

Our study revealed that prescriptions issued by neurologists (OR: 7.41 [1.21–45.22]) and cardiologists (OR: 6.20 [2.76–13.92]) were significantly associated with drug–drug interactions (p < 0.05).

## DISCUSSION

The frequency of drug–drug interactions (DDIs) observed in our study was 17.4%, which contrasts sharply with the findings of Allabi et al. (2020), who reported a 93% prevalence of DDIs among hospitalised patients in a cardiology unit in Benin [7]. This discrepancy may be explained by the fact that our study was conducted in community pharmacies, where prescribers may take the risk of combining drugs known to interact, while relying on effective clinical and paraclinical monitoring to manage potential adverse effects.

Cardiologists were twice as likely as other specialists to prescribe at least three medications. This may be attributed to the fact that cardiologists typically manage patients with multiple comorbidities and risk factors, often requiring combination therapy with several pharmacological agents.

The number of drugs prescribed was a significant risk factor for the occurrence of DDIs (p < 0.001), consistent with the findings of Keddad et al. [8]. This may be due to the presence of multiple interaction mechanisms: the more drugs a patient receives, the higher the likelihood of triggering one or more of these mechanisms, thereby increasing the overall risk of DDIs [4].

Cardiologists were six times more likely than other prescribers to generate prescriptions associated with drug–drug interactions. As previously discussed, cardiologists were also more likely to prescribe three or more medications per prescription, and the number of DDIs was significantly associated with the number of drugs prescribed. Consequently, the high risk of DDIs associated with cardiologists’ prescriptions appears to be the result of a logical sequence of prescribing practices.

Although neurologists were not statistically more likely to prescribe three or more drugs, they were seven times more likely to issue prescriptions involving DDIs than most other prescribers. This may be due to a high degree of similarity between neurology and cardiology patients in terms of medical histories and comorbidities, which often necessitate the use of complex pharmacological regimens.

## CONCLUSION

This study provided an overview of the factors generally favouring the occurrence of drug–drug interactions in medical prescriptions dispensed in community pharmacies, particularly in the Efoulan Health District of Yaoundé. Drug–drug interactions in the community pharmacy setting have a moderate frequency in Cameroon. They are statistically associated with the number of drugs prescribed as well as with certain prescriber specialties, notably cardiology and neurology.

## Data Availability

All data produced in the present study are available upon reasonable request to the authors

## REFERENCES

1. Nnanga N, Ngoule CC, Soppo V, Eyango MP, Mbole JM, Nkoa T. Evaluation of the Quality of Medical Prescriptions in Community Pharmacies in the 3rd District of Douala City. Health Sci Dis. 2018;19(4):26.

2. Locca JF. Quality of drug prescriptions: progress through physician-pharmacist collaboration. Rev Med Suisse. 2009.

3. Maida C. Drug interactions in the elderly: from theory to practice [Internet]. Doctoral thesis, Faculty of Pharmacy, Mohammed V University; 2008.

4. Becker ML, Kallewaard M, Caspers PW, Visser LE, Leufkens H, Stricker BH. Hospitalisations and emergency department visits due to drug–drug interactions: a literature review. Pharmacoepidemiol Drug Saf. 2007;16(6):641–651.

5. Gacem H, Beriala H, Hamzi A, Derghal R, Ahmane A. Drug interactions in clinical practice: a prospective study in a cardiology ward. Batna J Med Sci. 2014;1(1):2–6.

6. Nguele O, Bayaga H, Mballa N, Nnanga N. Evaluation of the prescription of psychotropic drugs and their potential drug interactions at the Jamot Hospital in Yaoundé, Cameroon. Int J Pharmacol Res. 2021;11(9):e5658.

7. Allabi AC, Tchabi Y, Hounkponou M, Quenum R, Vehounkpe-Sacca J. A prospective analysis of potential and observed drug–drug interactions, adverse events and associated risk factors in hospitalised cardiology patients in Benin. Curr Drug Saf. 2020;15(3):190–197.

8. Keddad A, Gacem H, Aissaoui A. Drug interactions in community pharmacies: retrospective analysis of 2801 prescriptions. J Med Sci. 2015;2(2):133–136.

